# A unique dexamethasone-dependent gene expression profile in the lungs of COVID-19 patients

**DOI:** 10.1101/2022.01.12.22269048

**Authors:** Ulrik Fahnøe, Andreas Ronit, Ronan M. G. Berg, Sofie E. Jørgensen, Trine H. Mogensen, Alexander P. Underwood, Troels K. H. Scheel, Jens Bukh, Ronni R. Plovsing

## Abstract

Local immunopathogenesis of COVID-19 acute respiratory distress syndrome (CARDS) and the effects of systemic dexamethasone (DXM) treatment on pulmonary immunity in COVID-19 remain insufficiently understood. To provide further insight into insight into immune regulatory mechanisms in the lungs of CARDS (with and without DXM treatment) and critically ill non-COVID-19 patients (without DXM treatment), transcriptomic RNA-seq analysis of bronchoalveolar lavage fluid (BALF) was performed in these patients. Functional analysis was performed using gene ontology and a blood transcription module, and gene expression of select pro-inflammatory cytokines, interferon-stimulated genes (ISGs) and auto-IFN antibodies were assessed. We found 550 and 2173 differentially expressed genes in patients with non-DXM-CARDS and DXM-CARDS, respectively. DXM-CARDS was characterized by upregulation of genes related to pulmonary innate and adaptive immunity, notably B-cell and complement pathway activation, antigen presentation, phagocytosis and FC-gamma receptor signalling. Pro-inflammatory genes were not differentially expressed in CARDS vs. non-COVID-19, nor did they differ according to DXM. Most ISGs were specifically upregulated in CARDS, particularly in non-DXM-CARDS. Auto-IFN autoantibodies were detectable in BALF of some CARDS patients. In conclusion, DXM treatment was not associated with regulation of pro-inflammatory pathways in CARDS but with regulation of other specific local innate and adaptive immune responses.

**summary:** This study identifies differentially expressed genes in bronchoalveolar fluid of COVID-19 acute respiratory distress patients with a distinct RNA expression profile of those treated with dexamethasone. These results challenge the concept of a COVID-19 specific cytokine storm.

## Introduction

Coronavirus disease 2019 (COVID-19) caused by severe acute respiratory syndrome coronavirus 2 (SARS-CoV-2) leads to distinct clinical manifestations, ranging from mild symptoms to severe disease, such as life-threatening acute respiratory distress syndrome (ARDS). COVID-19 ARDS (CARDS) is associated with a mortality rate of 40-50%,^1,2^ which may be even higher in patients requiring invasive mechanical ventilation.^2^

The understanding of the pulmonary SARS-CoV-2 immunopathology is limited by the fact that most studies have assessed systemic rather than lung immunity.^3^ Given that COVID-19 results from a direct pulmonary insult following invasion of the lower airways, it is particularly informative to study immune responses within the lungs. The few studies that characterized the immunological landscape at the cellular level in severe disease CARDS bronchoalveolar lavage fluid (BALF) reported prominent pulmonary immune cell invasion, most notably inflammatory myeloid cells,^4–7^ and expression of several pro-inflammatory mediators, of which interferon (IFN) responses have been proposed to be mechanistically linked to the severity of lung injury.^8^ However, a more detailed understanding of the specific mechanisms of COVID-19-mediated lung injury is pertinent to uncover how various immunomodulatory therapies, including dexamethasone (DXM),^9,10^ affect lung immunity.

In the present cross-sectional study, we investigated the transcriptomic profiles of the local pulmonary host responses during early-stage severe CARDS, non-COVID-19 ARDS and sepsis by per protocol BALF sampling early after intubation. Moreover, we compared the innate and adaptive immune transcriptomic profile of non- and DXM-treated CARDS patients to elucidate how this specific immune-targeted treatment modifies the pulmonary host defense, including the gene expression profiles of proinflammatory mediators and interferon stimulated genes (ISGs).

## Methods

### Study population, setting and ethics

Inclusion criteria of CARDS patients were age >18 years, SARS-CoV-2 infection confirmed by RT-PCR, presence of ARDS according to the Berlin criteria,^11^ and less than 72 hours of invasive mechanical ventilation. All patients were recruited during admission to the intensive care unit (ICU) at Hvidovre Hospital, Denmark (one of four ICUs in the Capital Region to which COVID-19 management was centralized). All patients were recruited between April 6^th^, 2020 and April 12^th^, 2021. This period included the first and second COVID-19 wave in Denmark, which corresponded to a time prior to and after DXM treatment was introduced as standard of care. Thus, patients recruited during the first wave did not receive DXM treatment. Supportive ICU treatment was not changed during this time period. We have previously presented flow cytometric results of 4 CARDS patients that were also included in the present study.^5^

As a comparator group, patients with ARDS from bacterial respiratory pathogens as well as mechanically ventilated patients with sepsis without ARDS were included; both of these patient groups were termed non-COVID-19. Except for SARS-CoV-2 infection, inclusion criteria were identical. All non-COVID-19 patients had ARDS and/or sepsis according to Berlin criteria^11^ and The Third International Consensus Definition for Sepsis and Septic Shock,^12^ respectively. Results from these patients have not been presented elsewhere. Finally, a healthy control group was included for comparison of IFN autoantibodies. Unrelated data from these healthy individuals have been presented elsewhere.^13,14^

All patients (CARDS and non-COVID-19) were sedated and unable to provide oral and written informed consent, which was therefore obtained from the next of kin.^5^ The study was approved by the Regional Ethics Committee of Copenhagen (H-20023159/H-22011021/H-22009131) and the Knowledge Center for Data Review of Copenhagen (P-2020-399) and registered at ClinicalTrials.gov (NCT04354584).

### Bronchoalveolar lavage procedure and sample processing

The BAL procedure was performed per protocol in the supine position on deeply sedated patients (adjuvant treatment strategy at the clinician’s discretion). In brief, FiO_2_ was increased to 1.0 if possible and the procedure was performed in a standardized fashion using a disposable videoscope with an outer diameter of 5.0 mm (Ambu aScope 4 Broncho Regular 5.0/2.2, Ambu A/S, Ballerup, Denmark). Three successive 50 mL aliquots of prewarmed (37°C) isotonic saline were instilled in the medial segment of the right middle lobe and aspirated immediately with low negative suction pressure (< 100 cm H_2_O). The duration of the BAL procedure was between 2 and 5 minutes and there were no procedure-related complications.

BALF was pooled into a sterile glass container on ice and processed within 15 min of collection. Pellet and fluid were separated using centrifugation at 1500 RPM for 7 minutes at 4°C. Pellet was resuspended with phosphate-buffered saline, centrifuged, and pellet was separated in two cryotubes with 600 μl undiluted ribonucleic acid (RNA)later (Sigma-Aldrich Co., Merck KGaA, Darmstadt, Germany) in each. Samples were stored at -80°C.

### Sample preparation and RNA extraction

For extraction, cells in RNAlater were thawed and pelleted by centrifugation. Pellet was resuspended in 750 μL TRIzol LS (ThermoFisher Scientific, Waltham, MA, US) and 250 μL RNAase free water was added. The lysate was transferred to a gel lock heavy tube and 200 μL chloroform was added. Tubes were shaken horizontally for 15 seconds and spun for 15 min at 12000 g at 4°C to allow phase separation. The aqueous phase was added to an equal volume of 100% EtOH and mixed. RNA extraction was performed on cc25 zymo column with an on-column DNase treatment following manufactures guidelines and eluted in RNase free water. RNA yield and integrity were assessed using Qubit (ThemoFisher Scientific) and Bioanalyzer 2100 (Agilent, Santa Clara, CA, US) using the RNA nano chip.

### Library prep and sequencing

100 ng RNA was used for each sample as input for library prep. The NEBNext rRNA Depletion Kit (Human/Mouse/Rat)(New England Biolabs, Ipswich, MA, US) was applied for ribosomal RNA depletion and NEBNext Ultra II Directional RNA Library Prep (New England Biolabs) was used together with dual index primers to generate libraries for sequencing. After assessment of library quantity and quality by Qubit and Bioanalyzer 2100, equimolar amounts of each library were pooled and run on two Nextseq 500 (Ilumina, San Diego, CA, US) v2 75 cycles kits to obtain 30-40 million reads per sample.

### Sequence analysis

Reads from the two flow cells were de-multiplexed, pooled and mapped by Hisat2 v2.1.0 to the human reference genome GRCh38 obtained from Ensembl. Gene expression profiles were obtained from the sorted bam files using featureCounts v2.0.0 and the Ensembl annotation GRCh38.103. Differential analysis was performed in R, as described previously.^15^ Briefly, we used the Limma-voom and DeSeq2 package for pre-processing and principal component (PC) analysis, respectively, and figures were produced using the ggplot2 package. The following four contrasts were compared: CARDS vs. non-COVID-19, DXM-CARDS vs. non-COVID-19, non-DXM-CARDS vs. non-COVID-19 and DXM-CARDS vs. non-DXM-CARDS. Gene ontology (GO) analysis was performed using Goseq on up and downregulated genes separately for each contrast. Analysis of ISGs was performed using ROAST on differentially expressed (DE) genes for each contrast using a curated list of 399 ISGs.^16^ Similarly, MROAST was applied to analyse enrichment of specific immune cell signatures as defined by the 2021 update of the blood transcriptome modules (BTMs; https://github.com/shuzhao-li/BTM), as described previously.^17^ Finally, we analysed inflammatory genes of interest, including interleukin (IL)-1A, IL-1B, IL-2, IL-6, IL-8/CXCL8, IL-11, IL-17A, IL-17B, IL-18, tumour necrosis factor (TNF)-α, IFN-λ, granulocyte-macrophage colony-stimulating factor (GM-CF/CSF2), granulocyte colony-stimulating factor GCSF/CSF3, monocyte chemoattractant protein-1 (MCP-1/CCL2), interferon gamma-induced protein (IP-10/CXCL10) and macrophage inflammatory protein-1 (MIP-1-α/CCL3), which were selected based on previous BALF-based studies.^5,6,18^ SARS-CoV-2 genome sequences were analysed as described previously.^19^

### IFN autoantibodies

IFN autoantibodies were measured in BALF by enzyme-linked immunosorbent assay (ELISA), as previously described for serum.^20^ Briefly, ELISA plates were coated with 1 μg/mL IFN-α (Miltenyi Biotec, Bergish Gladbach, Germany) and IFN-ω (ThermoFisher Scientific) overnight at 4°C followed by blocking in 5% skimmed milk. Bound autoantibodies were detected with HRP-conjugated goat anti-human IgG, IgA, IgM (Fc specific)(Nordic-MUbio, Susteren, Netherlands) and HRP substrate KPL SureBlue (Seracare Life Sciences, Milford, MA, US).

## Results

### Patient characteristics and bronchoalveolar lavage procedure

The BAL procedures in CARDS and critically ill non-COVID-19 patients were carried out after a mean of 28.8 hours (range 6-72) after intubation by the same physician (author RRP). All samples were obtained from the right middle lobe. A total of 13 CARDS patients and 8 non-COVID-19 patients were included (Table 1). All CARDS patients had moderate-to-severe impairment of oxygenation judged by the PaO_2_/FiO_2_ ratio and most (8/12) were treated with vasopressors at the time of the BAL procedure. Median duration of COVID-19 symptoms at the time of BAL procedure were 11 days (range 8-20). A total of 4/13 CARDS patients were not treated with DXM whereas 9/13 CARDS patients received DXM (6 mg per day for ten days) and 6/13 were also treated with remdesivir (all DXM treated). Non-COVID-19 patients encompassed 4/8 with ARDS and 4/8 with sepsis but without ARDS (Figure 1A). None of the patients with bacterial ARDS or sepsis were treated with corticosteroids.

**Table 1.** Clinical characteristics of patients with CARDS and non-COVID-19 patients.

| ID | COVID-19 | Sex | Age range# (years) | BMI (kg/m2) | ARDS* | PaO<sub>2</sub>/FiO<sub>2</sub> (mmHg)# | Vasopressor treatment | SAPS | Time from intubation to BAL procedure (hours) | Time from COVID-19 symptom onset to BAL procedure (days) | Outcome |
| --- | --- | --- | --- | --- | --- | --- | --- | --- | --- | --- | --- |
| C01 | No | M | 71-75 | 28 | Yes | 177 | No | 35 | 39 | NA | Survived |
| C02 | No | M | 66-70 | 24 | Yes | 115 | Yes | 61 | 43 | NA | Died |
| C03 | No | F | 66-70 | 22 | Yes | 175 | No | 44 | 13 | NA | Died |
| C04 | No | F | 61-65 | 17 | No | 112 | Yes | 45 | 20 | NA | Died |
| C05 | No | M | 61-65 | 31 | Yes | 113 | No | 36 | 10 | NA | Died |
| C06 | No | M | 35-40 | 25 | No | 199 | No | 38 | 39 | NA | Survived |
| C07 | No | M | 61-65 | 37 | No | 149 | Yes | 79 | 20 | NA | Survived |
| C08 | No | F | 61-65 | 27 | No | 427 | Yes | 81 | 36 | NA | Died |
| V01 | Yes | M | 41-45 | 21 | Yes | 83 | Yes | 58 | 65 | 10 | Survived |
| V02 | Yes | F | 61-65 | 25 | Yes | 108 | Yes | 72 | 44 | 9 | Died |
| V03 | Yes | M | 71-75 | 33 | Yes | 109 | Yes | 69 | 20 | 8 | Died |
| V04 | Yes | M | 71-75 | 26 | Yes | 70 | Yes | 77 | 72 | 20 | Survived |
| V05 | Yes (+DXM) | M | 55-60 | 22 | Yes | 115 | Yes | 56 | 41 | 14 | Survived |
| V06 | Yes (+DXM) | M | 71-75 | 29 | Yes | 112 | No | 54 | 10 | 17 | Died |
| V07 | Yes (+DXM) | M | 71-75 | 23 | Yes | 115 | Yes | 73 | 15 | 10 | Died |
| V08 | Yes (+DXM) | M | 56-60 | 31 | Yes | 123 | No | 56 | 6 | 12 | Survived |
| V09 | Yes (+DXM) | M | 66-70 | 25 | Yes | 150 | No | 55 | 12 | 11 | Died |
| V10 | Yes (+DXM) | M | 71-75 | 25 | Yes | 125 | Yes | 60 | 28 | 9 | Died |
| V11 | Yes (+DXM) | M | 66-70 | 26 | Yes | 86 | Yes | 59 | 18 | 14 | Survived |
| V12 | Yes (+DXM) | F | 45-50 | 16 | Yes | 100 | Yes | 58 | 24 | 20 | Died |
| V13 | Yes (+DXM) | F | 45-50 | 24 | Yes | 165 | No | 60 | 40 | 11 | Survived |

**Figure 1:**
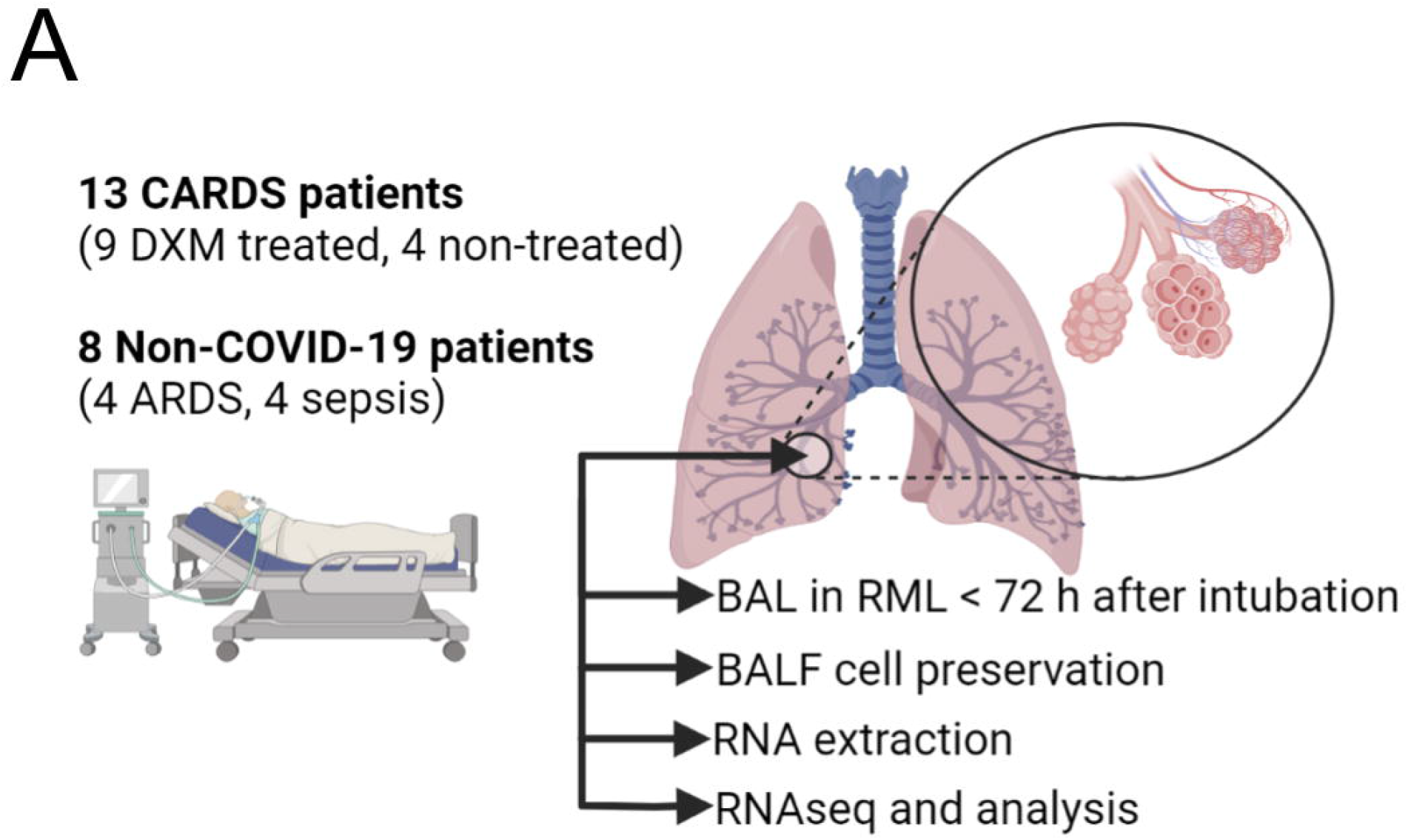
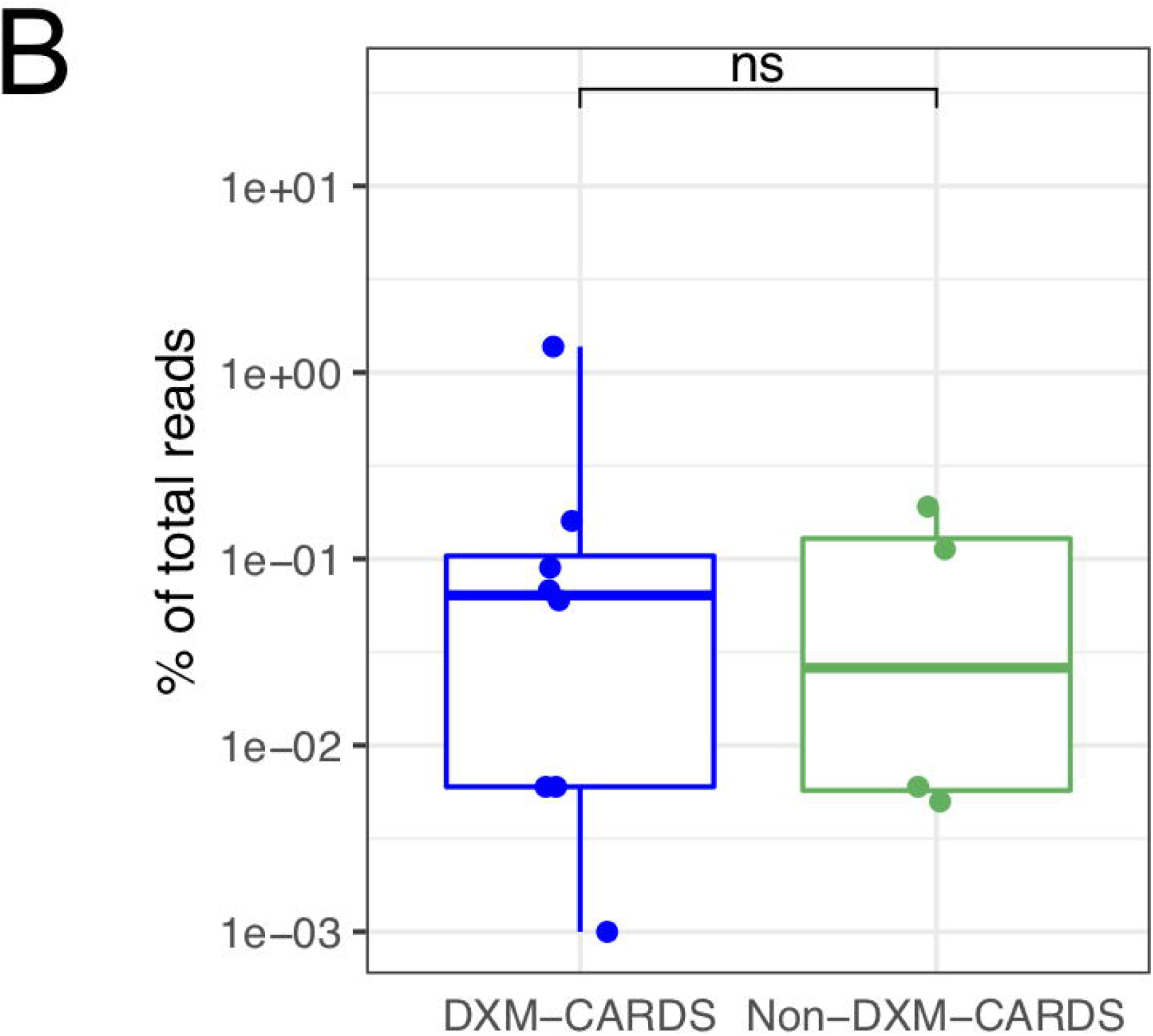
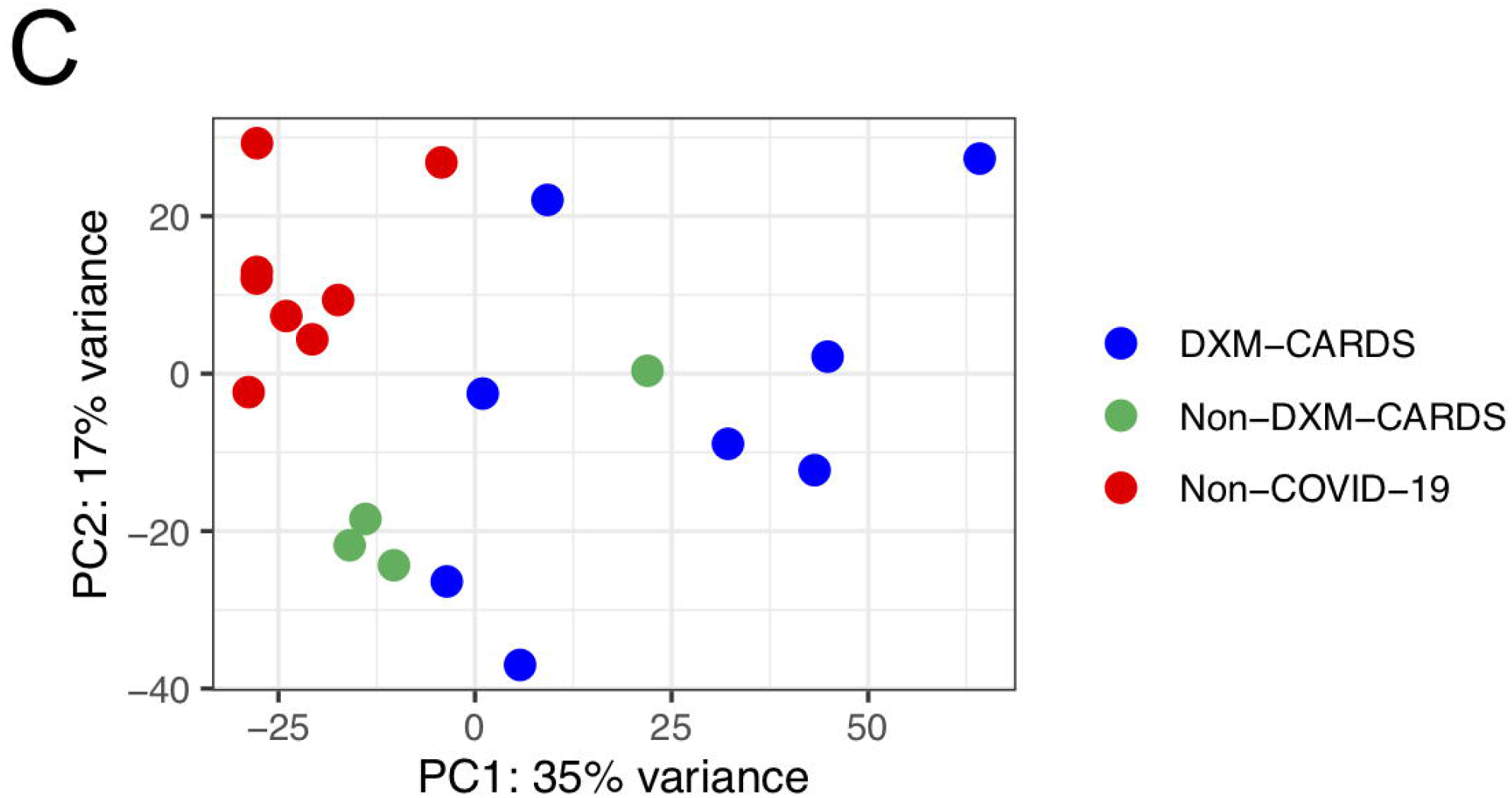
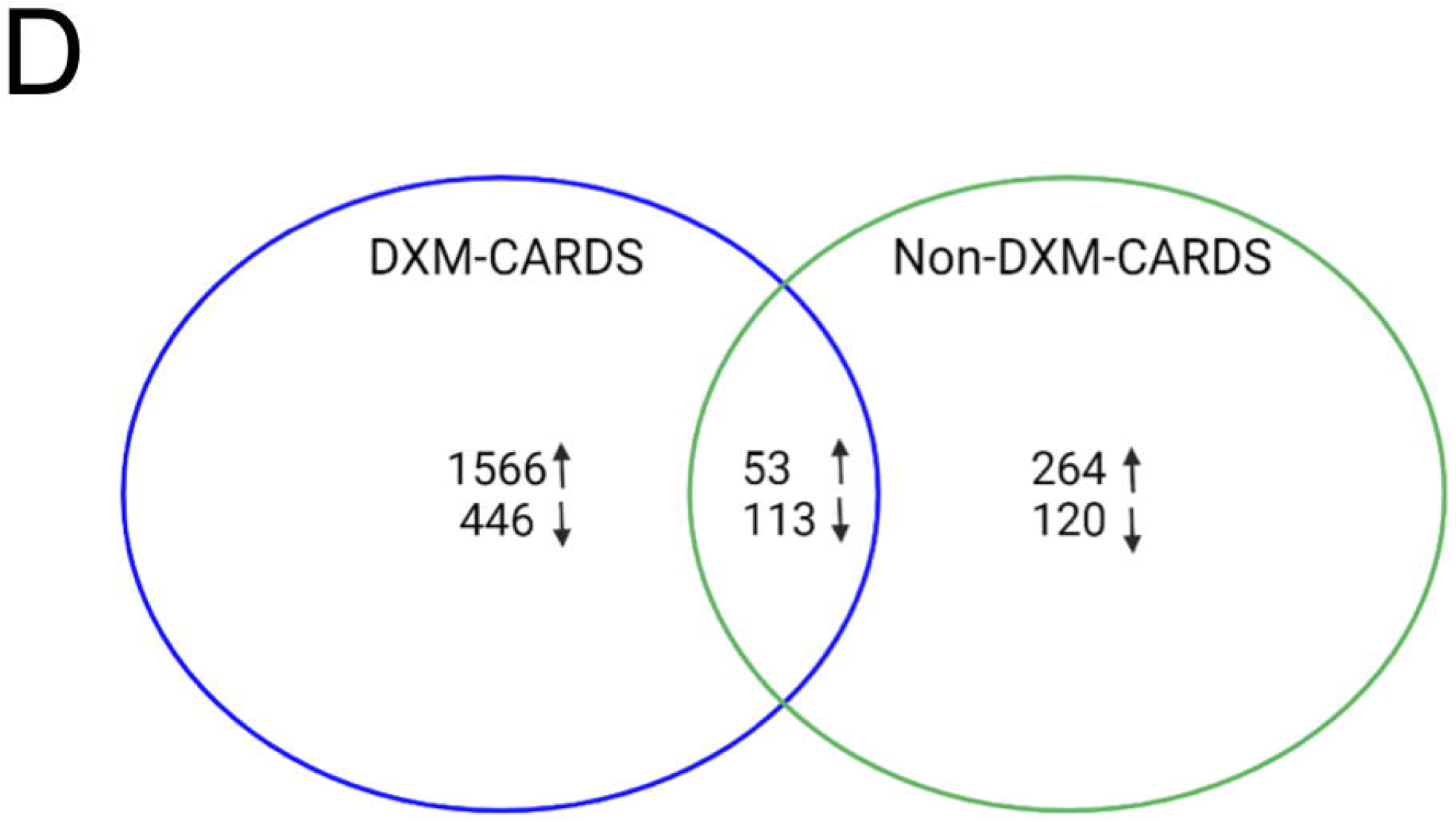
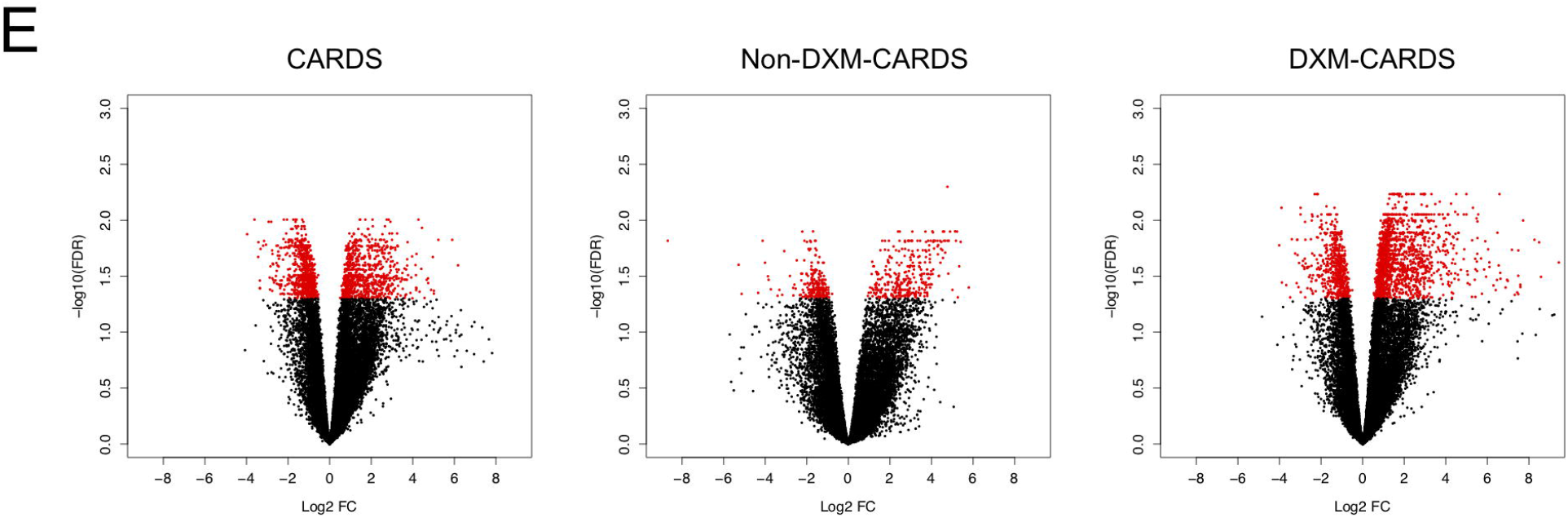
BALF transcriptome profile for CARDS patients. A) Experimental setup. B) Fraction (%) of viral reads (y-axis) compared to total number of reads shown as box plots. Lower and upper hinges correspond to the 25th and 75th percentiles. Differences analysed using Mann-Whitney *U* test. C) PC analysis depicting the three patient groups. D) Venn diagram representing the DE gene relationship for non-DXM-CARDS and DXM-CARDS compared to non-COVID-19. Arrows pointing up depict upregulated DE genes, and arrows pointing down depict downregulated DE genes. E) Volcano plots of the three comparisons; CARDS vs. non-COVID-19, non-DXM-CARDS vs. non-COVID-19, DXM-CARDS vs. non-COVID-19. DE genes for each plot are labelled in red. Abbreviations: BAL, bronchoalveolar lavage; CARDS, COVID-19 associated acute respiratory distress syndrome; DE, differentially expressed; DXM, dexamethasone; DE, differentially expressed; PC, principal component; RML, right middle lobe.

### Transcriptional features in the lungs of CARDS patients and during DXM treatment

Total RNA-seq was performed on cells obtained from BALF and the expression profile of CARDS and non-COVID-19 patients was compared. All except one patient (V05), in which there was not enough RNA, were included in the analysis. The levels of viral RNA in samples from CARDS patients varied greatly from 0.001-1.378% (Figure 1B). The seven samples with high fractions (above 0.1%) of viral reads showed differential coverage distribution skewed towards higher depth at the 3’ end of the viral genome compared to coverage at the 5’ end. This was concordant with the presence of sub-genomic RNA from replicating genomes indicating active infection of the cell. Furthermore, we found traces of negative strand RNA in those samples. In contrast, the five patients with a low fraction of viral RNA had more even coverage distribution over the entire genome suggesting limited replicating intra-cellular RNA.

PC analysis showed non-COVID-19 patients clustering closely together, while CARDS clustered more broadly but distinctly separate from the non-COVID-19 group (Figure 1C). Furthermore, non-DXM-CARDS clustered more closely than DXM-CARDS. Differential analysis of CARDS vs. non-COVID-19 revealed 852 up- and 827 down-regulated differentially expressed (DE) genes (false discovery rate; FDR < 0.05). Similar analysis performed with only non-DXM-CARDS revealed 317 up- and 233 downregulated DE genes, while DXM-CARDS had 1613 up- and 559 downregulated DE genes (Figure 1D-E). Additionally, we found 4 up- and 30 downregulated DE genes when comparing DXM-CARDS to non-DXM-CARDS. This indicated DXM as a primary driver of transcriptome changes in CARDS BALF.

### Functional signatures in the lungs of CARDS patients and during DXM treatment

Using GO analysis, we identified multiple cellular metabolic and rearrangement functions in addition to type-I IFN and T-cell activation represented by the upregulated genes during CARDS (Figures 2A and B), as well as enriched IFN I signalling. In addition, DXM-CARDS gene signatures were enriched for several B-cell activation and antigen presenting pathways, leukocyte migration and activation of complement. Interestingly, phagocytosis was also upregulated, especially FC-gamma receptor signalling. Only few downregulated gene categories were found, including lysozyme function, MHC class II responses and neutrophil degranulation.

**Figure 2:**
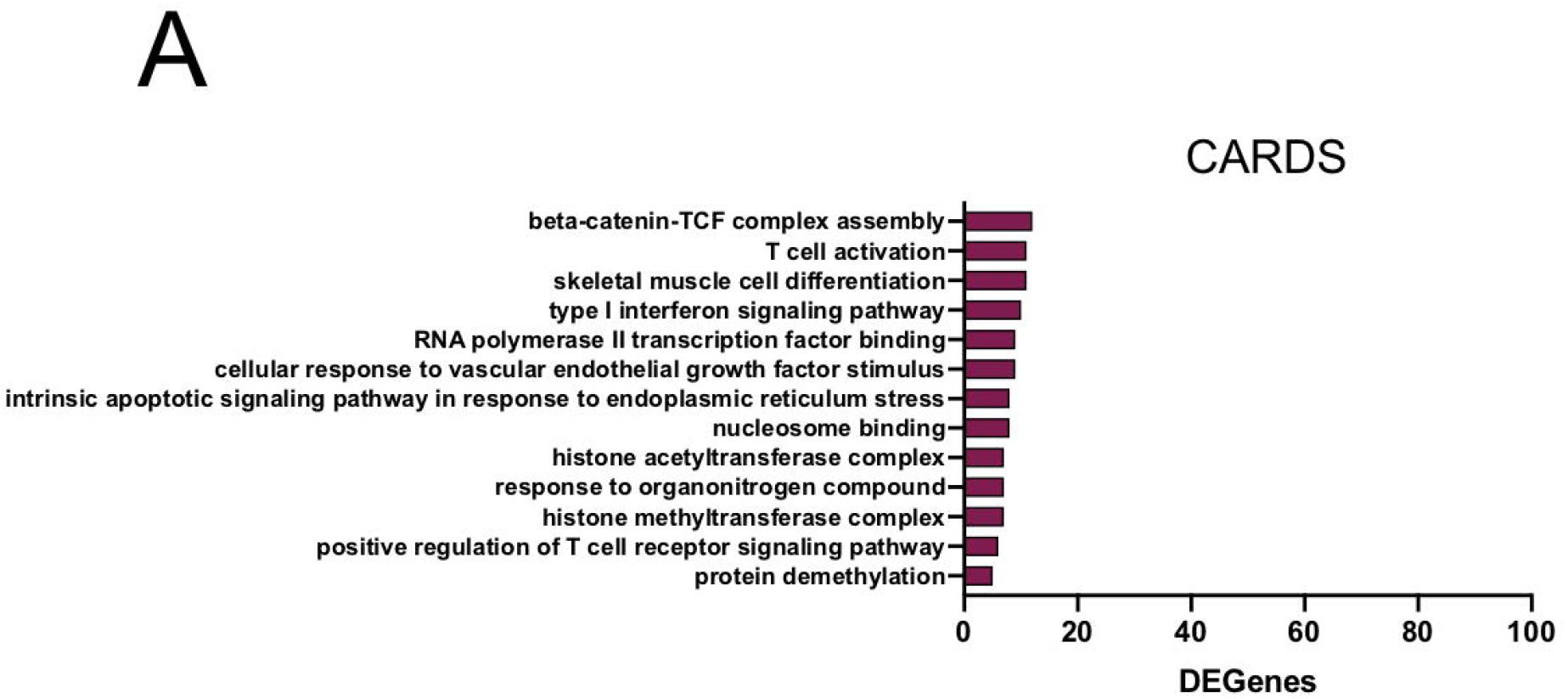
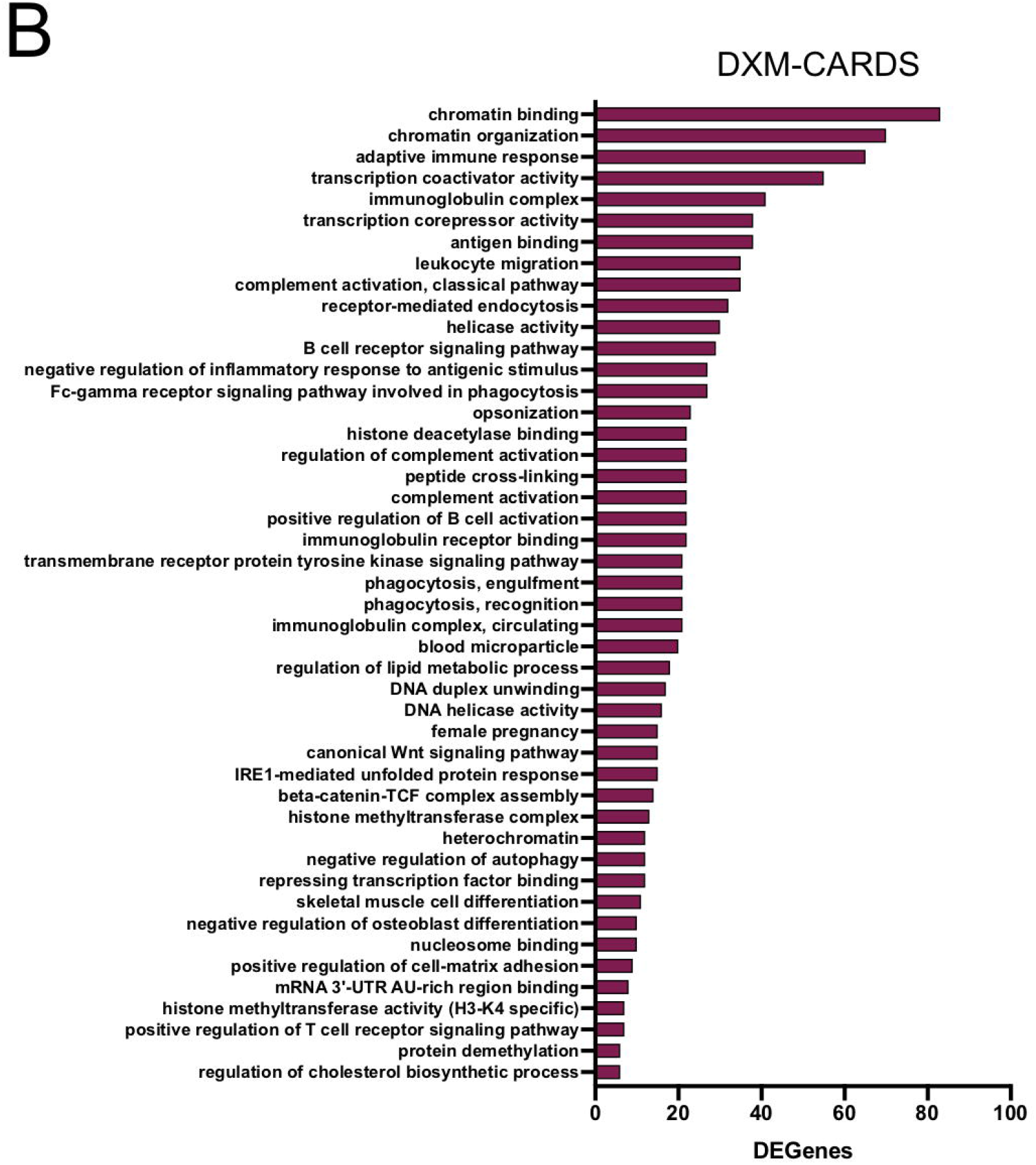
Gene ontology analysis in CARDS patients. GO analysis of upregulated DE genes for CARDS (A) and DXM-CARDS (B), respectively. Adjusted p-value set to p < 0.05 and only categories where DE genes accounts for more than 20% of total with 10 or more genes in the category displayed. X-axis shows the number of DE genes upregulated in each category. Abbreviations: CARDS, COVID-19 associated acute respiratory distress syndrome; DE, differentially expressed; DXM, dexamethasone.

To investigate immune cell responses more specifically, we intersected our transcriptomics data with BTMs, representing curated gene sets of correlated expression associated with biological function of circulating immune cells. We found significant upregulation of B-cell activation in the DXM-CARDS group (Figure 3), which is concordant with the GO analysis. In addition, CORO1A-def6, a modulator of interferon regulatory factor (IRF) 4 known to be involved in T-cell immunity and phagocytosis, was upregulated.

**Figure 3:**
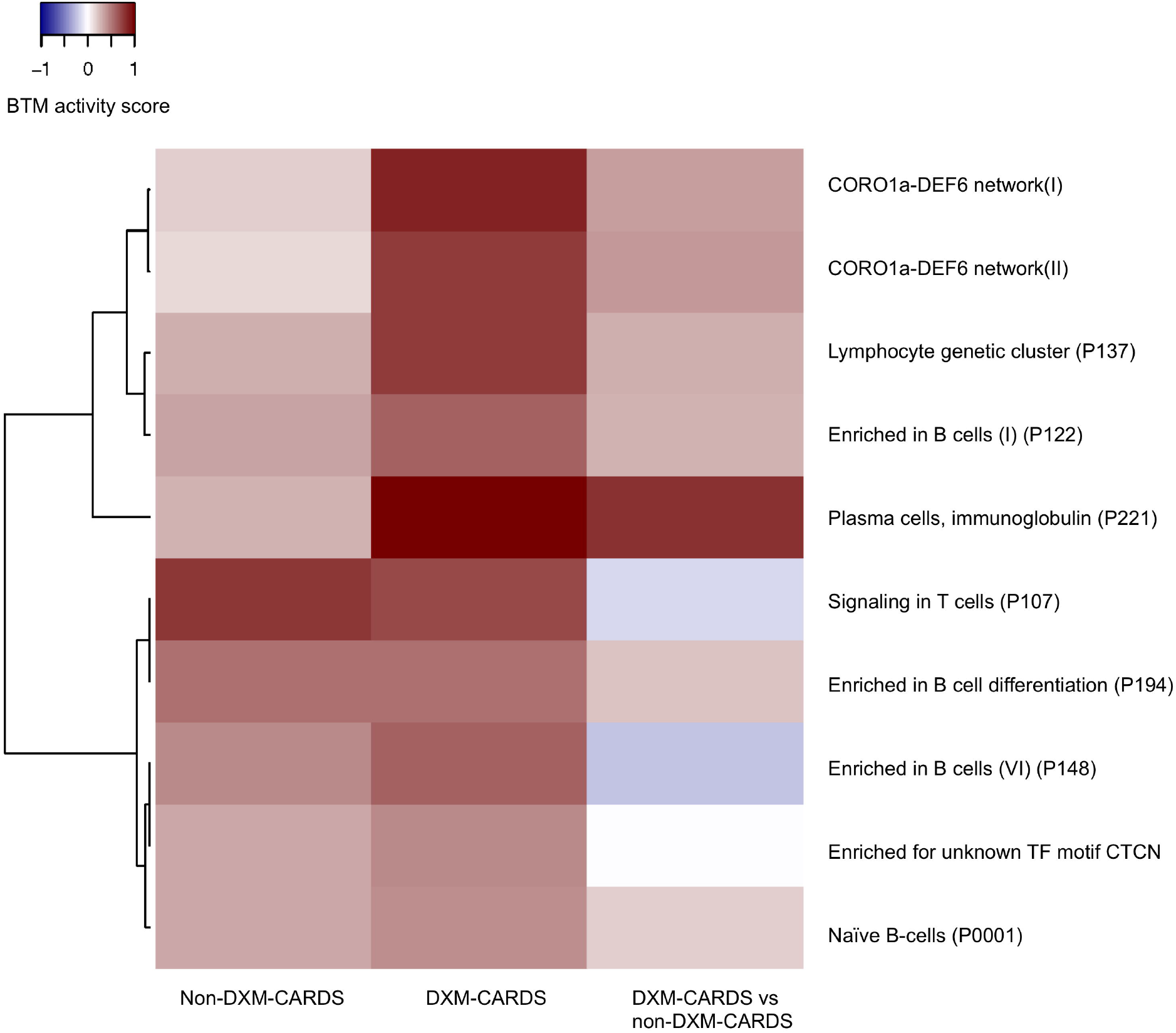
Heatmap of the BTM analysis depicting log2 fold-change level activity scores of BTM categories differentially enriched across all categories (MROAST p-value < 0.05). Non-DXM-CARDS vs. non-COVID-19 and DXM-CARDS vs. non-COVID-19 and DXM-CARDS vs. non-DXM-CARDS are represented in the columns. Rows represent the significant BTM categories. Abbreviations: BTM, blood transcription module; CARDS, COVID-19 associated acute respiratory distress syndrome; DE, differentially expressed; DXM, dexamethasone.

### IFN stimulated genes and IFN autoantibodies during DXM treatment in CARDS patients

We next assessed whether ISGs as a gene set was enriched in any of the conditions. Indeed, ISGs were enriched among upregulated genes, but specifically for non-DXM-CARDS patients (Figure 4A). Among genes differentially expressed across any condition (non-COVID-19, non-DXM-CARDS, DXM-CARDS), 42 ISGs were identified (Figure 4B). Most of these (36 genes) were elevated for CARDS and included typical responders to acute viral infection like ADAR, ATF3, IFITM3, IRF1, IRF7, ISG20 and TRIM56. A subset primarily enriched for DXM-CARDS included ABTB2, CNP, DDIT4, GALNT2, MAP3K14, PRKD2, SPSB1, SQLE, SSBP3 and TNK2. Curiously, six genes, FCGR1A, FKBP5, GLRX, LEPR, MSR1 and MT1L, were specifically upregulated for non-COVID-19 patients and may represent responses to bacterial infection. One non-COVID-19 patient (C07) had an ISG expression signature similar to CARDS patients, suggesting a putative undiagnosed viral infection in the that patient.

**Figure 4:**
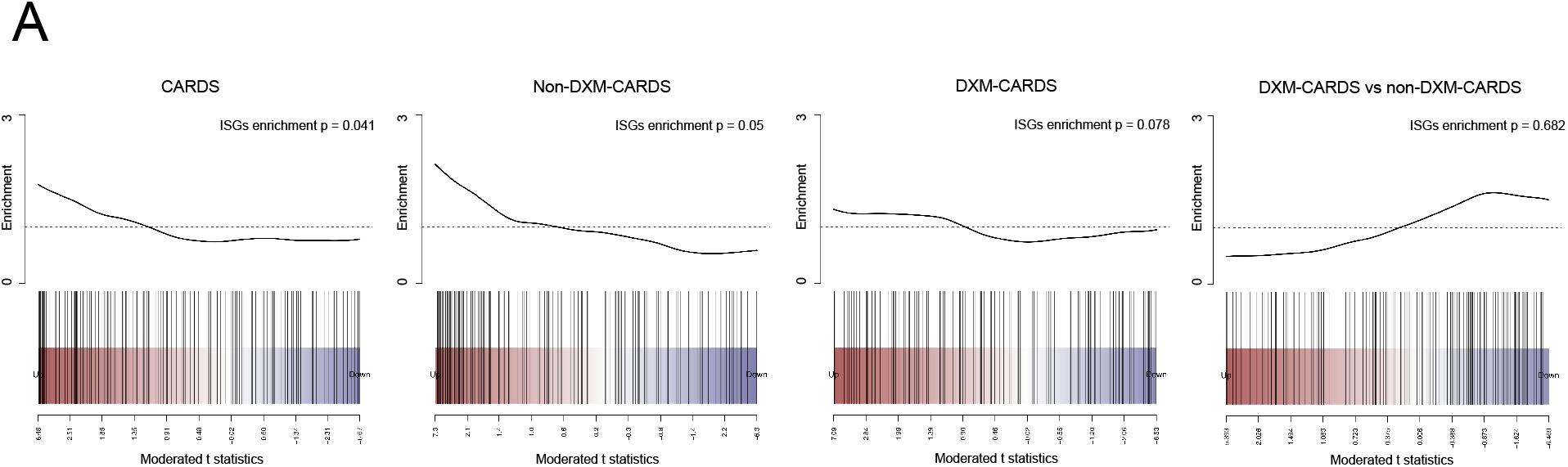
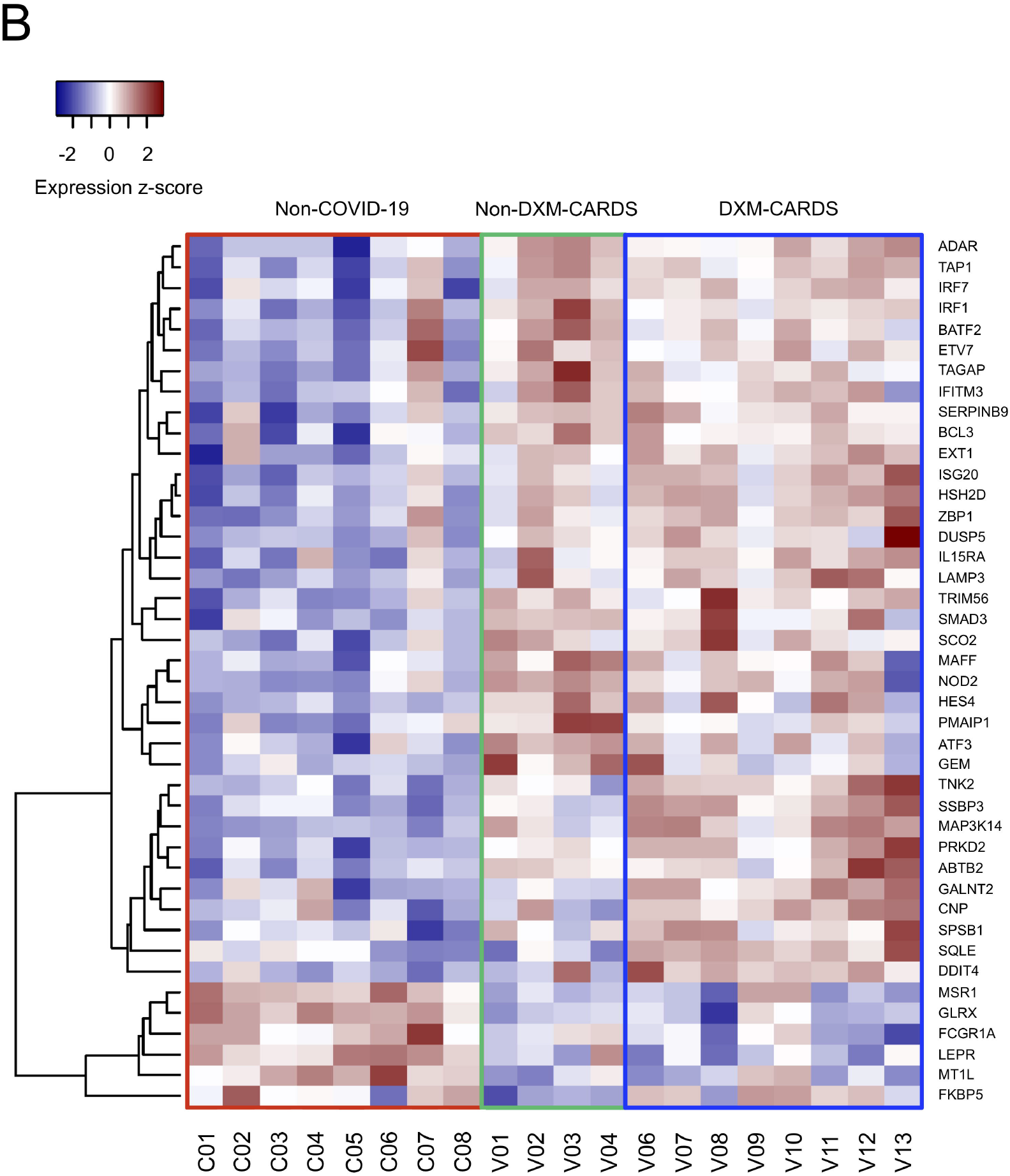
ISG regulation for CARDS patients. A) ISG enrichment analysis represented as barcode plots for contrasts CARDS vs. non-COVID-19, non-DXM-CARDS vs. non-COVID-19 and DXM-CARDS vs. non-COVID-19 and DXM-CARDS vs. non-DXM-CARDS. Enrichment analysis p-values from ROAST shown above each plot. Genes are ranked by differential expression statistics (moderated t statistic) on the x-axis with ISG gene set members depicted as bars. Corresponding line plot displays sliding average of set enrichment. B) Heatmap of differentially expressed ISGs showing expression z-scores. Rows represent individual ISGs and columns individual patients with patient IDs shown below. Boxes indicate patient groups: Non-COVID-19 (red), non-DXM-CARDS (green) and DXM-CARDS (blue). Abbreviations: CARDS, COVID-19 associated acute respiratory distress syndrome; DXM, dexamethasone; ISGs, interferon-stimulated genes.

IFN autoantibodies were measured in BALF from the 13 CARDS and 8 non-COVID-19 patients as well as 15 healthy controls (Figure 5A and B). We found elevated anti-IFN-α in DXM-CARDS patients compared to healthy controls, whereas anti-IFN-ω was elevated in both DXM-CARDS and non-DXM-CARDS. However, no differences between CARDS vs. non-COVID-19 patients were detected for anti-IFNα and anti-IFN-ω.

**Figure 5:**
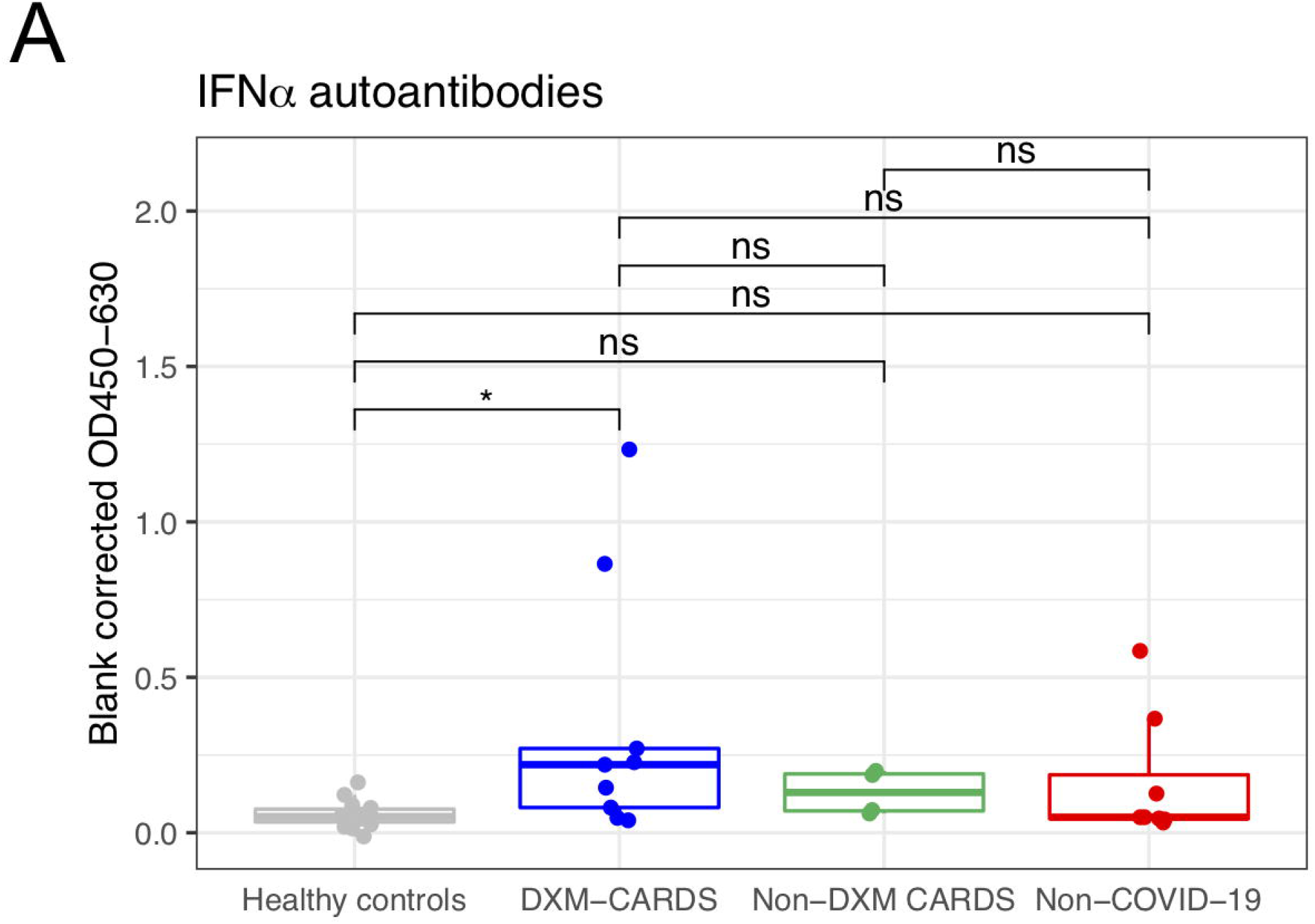
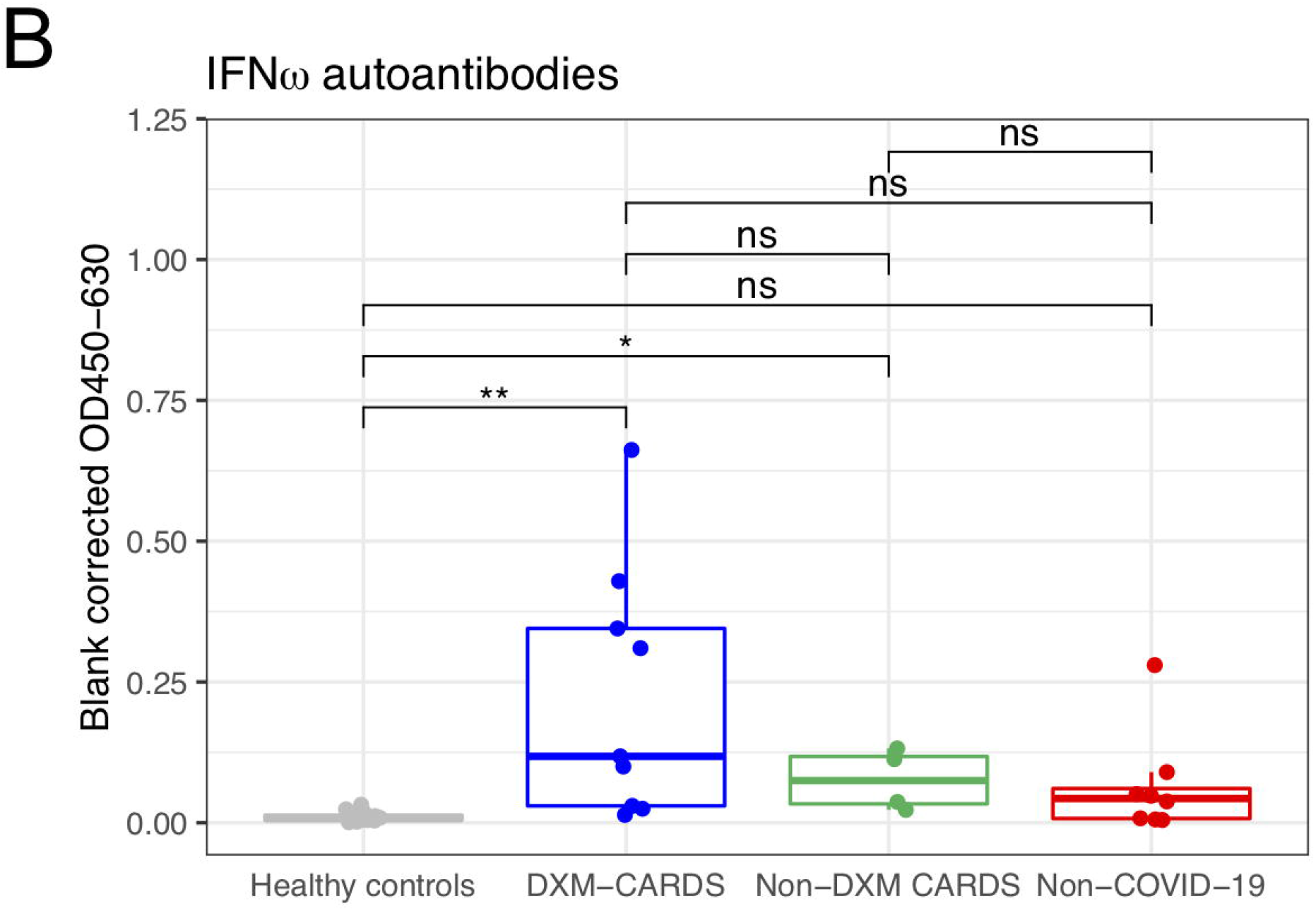
Autoantibodies against IFN shown for healthy controls, non-COVID-19, non-DXM-CARDS and DXM-CARDS patients. A) IFN-α autoantibodies and B) IFN-ω autoantibodies. Absorbance measured at OD450-630 corrected for blank measurement is shown on the y-axis. Lower and upper hinges of boxplots correspond to the 25th and 75th percentiles. Significant differences were analysed using Mann-Whitney *U* test. Abbreviations: CARDS, COVID-19 associated acute respiratory distress syndrome; DXM, dexamethasone; IFN, interferon; ISG, interferon stimulated gene.

### Inflammatory signatures of CARDS and during DXM treatment

When comparing cytokine/chemokine gene expression of IL-1A, IL-1B, IL-2, IL-6, IL-8/CXCL8, IL-11, IL-17A, IL-17B, IL-18, TNF, IFN-λ, GM-CF/CSF2, GCSF/CSF3, MCP-1/CCL2, IP-10/CXCL10 and MIP-1-α/CCL3, only the interleukin (IL)-18 gene was differentially expressed (downregulated) in CARDS compared to non-COVID-19. None of the genes were differentially regulated in DXM-CARDS vs. non-DXM-CARDS.

## Discussion

We identified differentially expressed genes in BALF of CARDS patients and a distinct RNA expression profile of DXM-treated CARDS patients that differed from non-treated patients. Functional analysis of DXM-CARDS revealed upregulation of cellular metabolism and rearrangement functions, enriched pathways for B-cell activation, antigen presentation and complement responses. ISGs were also particularly upregulated in non-treated CARDS patients. Contrary to our expectations, key pro-inflammatory genes were comparable for CARDS and non-COVID-19 and did not differ according to DXM treatment. Finally, we found evidence of IFN autoantibodies in the lungs of CARDS patients.

ARDS involves complex immunopathogenesis, which include influx of inflammatory cells (most notably neutrophils and macrophages), pro-inflammatory cytokines, proteases and procoagulant factors in the lungs.^21^ A local hyperactive immune response has also been noted in CARDS and the term “cytokine storm” has been widely used to describe its pathophysiology.^22^ However, studies have suggested that circulating levels of pro-inflammatory mediators in CARDS may be lower than in patients with other causes of ARDS.^23^ We here aimed to identify specific local mechanisms of COVID-19 mediated lung injury by comparing the pulmonary transcriptional profile of CARDS and non-COVID-19 patients. Although we and others have previously shown a high expression of inflammatory cytokines and chemokines (i.e. neutrophil chemotactic factor IL-8, IL-6, CCL2/MCP-1, IFN-γ-inducible protein 10 IP-10/CXCL10 amongst other) in CARDS patients compared to healthy controls,^5,6,24^ we did not find a differential expression when compared to critically ill non-COVID-19. However, here we did find a total of 1613 and 317 other DE genes in DXM-CARDS and non-DXM-CARDS patients, respectively, of which only 53 were upregulated in both groups, compared to non-COVID-19 patients. Although glucocorticoid treatment is known to suppress the production of at least some inflammatory cytokines in other diseases,^25,26^ and has indeed been posited to improve outcome in CARDS by suppressing the “cytokine storm”,^27^ we did not observe a differential gene expression according to DXM treatment of pro-inflammatory mediators in the lungs.

Clinical trials have provided evidence that treatment with DXM at a dose of 6 or 12 mg per day for up to 10 days reduces 28-day mortality in patients hospitalized with COVID-19.^9,28^ Glucocorticoid treatment has on the other hand also been associated with increased mortality in other infectious diseases, including influenza.^29^ This discrepancy suggests a COVID-19 specific response profile that renders DXM effective, and it has been hypothesized that COVID-19 may induce glucocorticoid insensitivity in the process of modulating host cell activities.^30^ In our GO analysis, complement responses, particularly genes related to the classical pathway initiated by antigen-antibody complexes, were specifically upregulated in DXM-CARDS. Complement activation is an important component of the innate immune response to viral infections ^31^ and has been associated with ARDS pathology of other causes.^32^ However, the clinical role of local complement activation in CARDS remains insufficiently understood. One study found increased levels of soluble C5a correlating with severity of COVID-19 with high expression levels of C5aR1 receptors in blood and pulmonary myeloid cells.^18^ Why complement activation was preferentially upregulated in DXM-CARDS patients is unknown. In contrast to this, *in-vitro* and *in vivo* studies unrelated to COVID-19 have shown reduced local complement activation following DXM treatment.^33,34^ In addition, we found enriched B-cell signatures in GO analysis and BTM analysis. Early studies performing flow cytometry and single cell RNA-seq found low proportions of B-cells in BALF of COVID-19 patients,^4,5^ whereas another RNA-seq study, using a deconvolution analysis, revealed higher proportions of B-cells in BALF compared to healthy controls,^35^ thus suggesting that DXM preferentially stimulates B-cell immunity to SARS-CoV-2. In line with this, early studies have revealed a direct effect of glucocorticoids on human B-cell survival and development.^36,37^ We also found upregulation of the phagocytic pathway especially FC-gamma receptor signalling, which is associated with antibody-bound viruses and concomitant phagocytosis.^38^ In addition, the BTM analysis found the CORO1A-DEF6 network, linked to T-cell and phagocytic function,^39^ to be enriched, which has also been observed for influenza vaccination.^40^

Type-I IFNs (e.g., IFN-α and IFN-β) provide protective immunity against SARS-CoV-2 as highlighted by studies showing that inborn errors of TLR3- and TLR7-dependent antiviral innate immune pathways may underlie severe COVID-19.^41^ Previous *in vitro* studies of other viral infections have shown that glucocorticoids may suppress type-I IFN-mediated responses by inhibiting intracellular signalling pathways and subsequent expression of ISGs.^42,43^ This correlated well with our observation of enrichment of ISGs specifically in the BALF of non-DXM-treated CARDS patients. In addition, we found one distinct cluster of ISGs to be upregulated in DXM-treated patients (TNK2, SSBP3, MAP3K14, PPKD2, ABTB2, GALNT2, CNP, SPSB1, SQLE, DDIT4) and one cluster of downregulated ISGs for all CARDS patients (MSR1, GLRX, FCGR1A, LEPR, MT1L, FKBP5).

IFN-α and IFN-ω autoantibodies circulating in blood or present in tissue fluids/compartments can neutralize the ability of type-I IFNs to block viral infection. They have been identified in serum of ∼10-15% of patients with critical COVID-19 pneumonia and in around 20% of all deaths from critical COVID-19 across all age groups, whereas they are typically absent (or present in a minor fraction, i.e. 0.5%) in individuals with asymptomatic or mild disease.^20,44^ Moreover, IFN autoantibodies have been found to be present from early time points (4-13 days) in the course of COVID-19.^44,45^ Blank corrected serum levels of > 0.5 are usually considered clinically relevant and sufficient for neutralization.^20,46^ We here identify a significantly higher level of antibodies towards IFN-α and IFN-ω in the lungs of CARDS patients, where they may negatively impact local antiviral immunity to SARS-CoV-2. To our knowledge, the presence of IFN autoantibodies have not previously been assessed in BALF of patients with CARDS or in any other group of mechanically ventilated patients. Future studies with larger sample sizes should correlate BALF IFN autoantibody levels with SARS-CoV-2 viral load and ISG responses to define the role of these autoantibodies on COVID-19 lung pathophysiology

In contrast to prior COVID-19 studies assessing the local pulmonary immune response, in which BALF was primarily collected on clinical indication at a later stage, and at the clinician’s discretion regarding site and volume of lavage, a strength of our study is the per protocol BAL sampling in the early course of severe disease progression using a uniform cross-sectional approach. Although we were able to identify local immune signatures of CARDS patients and specific DXM treatment effects, our sample size is small and results should be confirmed in larger and independent studies with several time points.

The complexity of the pulmonary host responses to pathogen invasion is well known and for CARDS both innate and adaptive immune responses play crucial roles in containing and resolving infection. Our results provide further insight into the pulmonary host response of CARDS and potential effects of DXM treatment but they also raise several important questions and specifically challenge the concept of a local “cytokine storm” as a driver of respiratory failure during COVID-19.

## Data Availability

The following sentence was used in the manuscript: "All data produced in the present study are available upon reasonable request to the authors" 

## Abbreviations

(CARDS): COVID-19 acute respiratory distress syndrome
(BALF): bronchoalveolar lavage fluid
(DXM): dexamethasone
(COVID-19): coronavirus disease 2019
(SARS-CoV-2): severe acute respiratory syndrome coronavirus 2
(ARDS): acute respiratory distress syndrome
(IFN): interferon
(ISGs): interferon stimulated genes
(ICU): intensive care unit
(GO): gene ontology
(DE): differentially expressed
(BTMs): blood transcriptome modules
(TNF): tumour necrosis factor
(GM-CF/CSF2): granulocyte-macrophage colony-stimulating factor
(MCP-1/CCL2): monocyte chemoattractant protein-1
(IP-10/CXCL10): interferon gamma-induced protein
(MIP-1-α/CCL3): macrophage inflammatory protein-1
(ELISA): enzyme-linked immunosorbent assay
(IRF): interferon regulatory factor
(IL): interleukin

## Acknowledgments

We thank all the patients and their families for their participation. We thank Anna Louise Sørensen for her outstanding technical laboratory assistance. We thank the clinical staff and nurses at the Department of Anesthesiology and Intensive Care for their dedicated contribution.

